# MOSAIC (MOthers’ AdvocateS In the Community) for Pregnant Women and Mothers of Children Under 5 with Experience of Intimate Partner Violence: A pilot randomized trial study protocol

**DOI:** 10.1101/2022.04.14.22273885

**Authors:** Maji Hailemariam, Caron Zlotnick, Angela Taft, Jennifer E. Johnson

## Abstract

**Background:** Pregnancy and motherhood increase the risk for long-term exposure to physical, psychological and sexual intimate partner violence (IPV; sexual or physical violence by current or former partners). Pregnant women and mothers with children under 5 who have experienced IPV exhibit poor physical and mental health and obstetric outcomes. Depression and posttraumatic stress disorder (PTSD) are the two most common mental health consequences of IPV. There is good evidence that women with good social support have better mental health and IPV outcomes.

**Methods:** This study will develop MOthers’ AdvocateS In the Community (MOSAIC) Plus intervention for pregnant women and mothers with children under the age of 5. MOSAIC uses trained mentor mothers and has been found to reduce subsequent IPV. This study will blend the original MOSAIC intervention with principles of interpersonal psychotherapy (IPT) to address symptoms of depression, PTSD, and prevent subsequent risk of IPV. We will conduct a pilot randomized trial of the MOSAIC Plus intervention compared to the traditional MOSAIC intervention to determine its feasibility and acceptability. Study samples include focus groups (n=36), open trial (n=15), and a randomized pilot trial including 40 pregnant women and mothers with children under 5 who report current/recent of IPV and elevated symptoms of maternal depression and/or PTSD. The study’s primary outcome will be changes in maternal depressive and PTSD symptoms. Secondary outcomes will include reduction in subsequent IPV, improvement in functioning, changes in social support and effectiveness in obtaining resources.

**Discussion:** This is a formative study evaluating the feasibility and acceptability of a mentor mother intervention for pregnant women and mothers with children under 5. Promising results of this study will be used for a larger, fully-powered randomized trial evaluating the effectiveness of a mentor mother intervention in preventing subsequent IPV and reducing depressive and PTSD symptoms in this population.

**Strengths and limitations of this study:**

- The study is informed by a robust qualitative approach to intervention development that involves a series of focus group discussions.
- This study aims to develop an intervention that reduces future intimate partner violence, while also addressing related maternal mental health outcomes.
- A rigorous and reproducible design includes randomization, clear inclusion criteria, manualized treatment protocols and fidelity assessments.
- The study will use reliable and validated measures.
- Given the small sample size, results from the pilot randomized trial are underpowered to draw firm conclusions about effectiveness.

## Background

Violence against women is a global human rights concern and a significant public health challenge; one in three women experience lifetime physical and/or sexual violence.(1) As many as 5.3 million U.S. women experience intimate partner violence (IPV; referring to sexual or physical violence by current or former partners) annually(2). Depression and posttraumatic stress disorder (PTSD) are the two most common mental health consequences of IPV. In turn, developing depressive and PTSD symptoms after experiencing IPV increases future risk of IPV(3). IPV is also associated with chronic physical health problems(4, 5) and self-harm (6).

Pregnancy and motherhood increase the risk for prolonged exposure to physical, psychological and sexual IPV(7). IPV can start or worsen during the perinatal period. As many as 4% to 8% of women report IPV during pregnancy (8). Compared to those who do not experience IPV, mothers who experience IPV during pregnancy or in the year following childbirth are at elevated risk for experiencing physical injury, other medical challenges, homicide(9) and mental health problems including suicide(10, 11). Twenty percent of postpartum deaths are attributable to suicide(12).

Pregnant women and mothers who experience IPV may not seek care when they need it(13). For example, perinatal women with experience of IPV may not initiate prenatal care until third trimester(14). They are more likely than women without IPV to miss three or more prenatal visits(15). Perinatal women with experience of IPV are twice as likely to have babies with low birthweight(16), higher rates of severe acute maternal morbidity(17) and have infants requiring intensive care(18). IPV increases risk of depressive and PTSD symptoms, which in turn increases risk of subsequent IPV(19-21) (8, 21).

Interventions are needed that address both, especially for pregnant women and mothers of young children.(22-24). There are very few interventions that effectively reduce IPV among pregnant women and mothers with children under 5 who report IPV(22-24). One of them, Mother AdvocateS In the Community (MOSAIC) forms the basis of this study(25). Very few existing interventions address maternal depressive and PTSD symptoms that often follow IPV (3, 26, 27). An integrated intervention that addresses elevated symptoms of maternal depression and PTSD while reducing subsequent IPV is needed.

Mother AdvocateS In the Community (MOSAIC)(25) is a non-professional intervention delivered by mentor mothers from the community to reduce IPV in pregnant women and mothers with children under 5(28). The intervention combines elements of mentoring and IPV-specific support provided by mentor mothers. MOSAIC was tested in a fully-powered RCT for pregnant women and mothers with children under 5 in Australia and was found to reduce subsequent IPV(24). However, the effects of the intervention depression were not significant and PTSD symptoms were not evaluated. The proposed study will augment MOSAIC with principles of an evidence-based intervention to improve maternal mental health. Interpersonal psychotherapy (IPT) is the front-line treatment for perinatal depression(29) and has been found to reduce PTSD symptoms in the perinatal period(30). IPT addresses maternal mental health by helping women increase their general social support systems and build communication skills and confidence to access needed resources and help. IPT can be effectively delivered by lay providers(31). IPT is theoretically consistent with the MOSAIC approach and addresses the consequences of IPV. IPT focuses on building or better utilizing one’s social support network within the context of a life stressor (32, 33). The proposed study aims to integrate IPT principles into MOSAIC to address both IPV and its mental health sequelae.

Specifically, the study aims to:

1. Develop and pilot-test the feasibility and acceptability of MOSAIC Plus intervention in a sample of 40 women.
2. Evaluate changes in symptoms of depression and PTSD.
3. Assess reduction in subsequent IPV.
4. Measure changes in functioning, self-care, general health and wellbeing, and
5. Evaluate changes in social support and improvements in effectiveness obtaining resources.

## 2. Methods and analysis

### 2.1. Development Phase

#### 2.1.1. Project advisory board

We will establish a project advisory board that will include 6-8 pregnant women and mothers with children under 5 and potential mentor mothers from the community. They will provide feedback on the content of the intervention in the development stage and on study procedures and results generally.

##### Manual development

The goal of the development phase will be to develop the MOSAIC Plus intervention manual. We will begin with the MOSAIC intervention manual which was developed to reduce subsequent IPV experiences in perinatal women and mothers with children under 5 (28)but was not specifically designed to address concomitant mental health issues. The MOSAIC intervention uses non-professional befriending, domestic violence-specific advocacy and mentoring by mentor mothers from the community(34). The mentor mothers may or may not have lived-experience with IPV, maternal depression or PTSD. MOSAIC Plus will expand the social support offered by mentor mothers in MOSAIC with IPT approaches for broadening women’s social support network generally. In MOSAIC Plus, we will expand the existing MOSAIC manual by integrating IPT approaches to address depressive and PTSD symptoms among IPV-exposed mothers. IPT conceptualizes interpersonal functioning and disturbance in social support as intimately linked to psychological symptoms(35). IPT identifies a difficult life event or current interpersonal problem and addresses it by helping the individual to build or better utilize the individual’s extended social support network with specific IPT strategies that include psychoeducation, communication skills practice, and exploration/encouragement.

##### Focus group discussions

We will conduct focus group discussions with potential study participants and potential mentor mothers to enhance and tailor the MOSAIC intervention to address mental health outcomes (reduction in depressive symptoms and PTSD symptoms). We will use information generated from focus group discussions to ensure that our manual expansion optimally addresses the needs of our target population, the mentor mothers, and the eventual implementing systems. We will conduct six focus group discussions of 6-8 members each (for 30-44 in total). Three groups with pregnant women and mothers of young children with current, recent or prior experience of IPV, 2 groups with potential mentor mothers, and one group with staff from community agencies (the settings where we would envision MOSAIC Plus being implemented if found promising). Focus group discussions will explore perspectives of potential clients and mentor mothers regarding the potential acceptability, benefits, and risks of the proposed MOSAIC Plus intervention and of the IPT components we plan to integrate. The goal of the 3 focus groups with pregnant women and mothers with children under 5 with IPV experiences will be to explore: (1) women’s perspectives on the greatest needs of IPV-exposed perinatal mothers; (2) what they would like most from mentor mothers to help with IPV, depression, and PTSD symptoms; and (3) feedback on the proposed MOSAIC Plus intervention and on the IPT-based components to be integrated. The goal of the two focus groups with nurse-family partnerships will be to explore ways to maximize acceptability of MOSAIC Plus, to integrate the intervention into their ongoing programs, and how to design the MOSAIC Plus program to maximize future implementability.

Participants for potential clients focus group discussions will be recruited from Hurley Medical Center’s perinatal clinic and YWCA using our standard study recruiting procedures and criteria. For the other two focus group discussions, we will recruit participants from local agencies serving mothers.

##### Data analysis and manual development

A moderator accompanied by a note taker will facilitate the focus group discussions, which will be recorded by using a digital audio recorder. The recordings will be transcribed verbatim by an experienced transcriber. We will use the framework approach by using a framework drawn from the data. We will modify our coding framework based on the newly emerging themes. For rigorous and high-quality data management, we will use NVivo qualitative analysis software, version 12(36). We will develop an outline of the MOSAIC Plus intervention manual based on the original MOSAIC intervention manual. We will embed contents relevant to reducing depressive and PTSD symptoms to the new manual based on results of the focus group discussions. We will enhance, refine and adapt the MOSAIC Plus intervention for the outcomes of interest based on the results of the focus group discussions. Dr. Hailemariam will conduct member checking with selected study participants to validate the fit, credibility and transferability of the results (i.e., main themes and translation of them into the manual itself) from the focus group discussions.

##### Open trial

The second part of the development phase will include conducting a small open trial to further refine the intervention manual and assess feasibility of research procedures. We will recruit 15 pregnant women and mothers with children under 5 with experience of IPV, who also report elevated symptoms of depression and/or PTSD, meeting the same inclusion criteria as participants in the randomized trial (see sampling and recruitment section below), and who will receive the MOSAIC Plus intervention. We will review our strategies for recruitment and retention of participants, strategies for recruitment, supervision and retention of the mentor mothers and mentor mothers’ compliance with the study protocol. Participants will be requested to complete an intervention specific End-of-Treatment Questionnaire assessing perceived helpfulness of various components of the intervention and their comfort with the research processes and assessments. We will review the responses of participants at an exit interview to elicit information for further refining the intervention protocol.

##### Program and procedure modifications

Based on data from the development phase, exit interviews, and ongoing feedback from the study team, the intervention manual, fidelity scales and the training program will be modified. Study recruitment and retention procedures will also be revised based on experiences of the research team and participants. These procedures will enhance study feasibility by ensuring that our assessments do not place undue burden on participants that measures are feasible and acceptable; and that dropout rates are minimized.

### Pilot Study Phase

In the **pilot study phase**, the MOSAIC Plus intervention will be compared to the traditional MOSAIC in 40 women. The goal of this phase will be to assess the feasibility of the research design, the acceptability of the MOSIAC Plus intervention and to explore 95% confidence intervals of reduction around depressive and PTSD symptoms, and decrease in the experience of IPV within a 9-month period from intake. The duration of both interventions will be 9 months. Study assessments will take place at baseline, 3, 6 and 9 months.

#### Sampling and recruitment

The randomized pilot study will involve 40 women who are 1) pregnant and/or are mothers of children under 5 and report IPV experiences in the past 6 months (as assessed by Composite Abuse Scale(37)) 2) aged 18 or above, and 3) have elevated depressive and/or PTSD symptoms (including full disorder). This will be determined using the Patient Health Questionnaire (PHQ-9) with a cutoff point of ≥9(38), and the Davidson Trauma Scale (DTS) with a cut-off point of ≥40(39). We anticipate that the majority of study participants will have both elevated depressive and PTSD symptoms, given that 50% of individuals with PTSD also have comorbid major depressive disorders, (40, 41) especially among women with IPV(21). Participants will be excluded if they (1) cannot provide the name and contact information of at least two locator persons (∼6%), and/or (2) do not have access to any telephone, (3) have a current serious mental illness requiring intensive care or (4) cannot understand English well enough to understand the consent form or assessment instruments when they are read aloud. Virtually all pregnant women and mothers of children under 5 accessing care in these settings speak English. An initial sample of 15 pregnant women and mothers with children under 5 will be recruited to participate in a nonrandomized open pilot trial and will receive the proposed MOSAIC Plus mentoring intervention. No woman will be excluded from recruitment on the basis of race, sexual orientation, gender identity? or disability status.

Participants will be recruited from two local agencies, the Hurley Medical Center OBGYN clinic and the YWCA-Flint. The YWCA offers domestic violence support, emergency shelter and long-term housing for persons with domestic violence exposure. For women identified as having a positive response to screening questions at those agencies, the provider in charge will ask “would you be interested in meeting with a person to hear about a voluntary project about peer support for pregnant women and mothers?”. The assessing provider will also explain that their decision whether to obtain additional information about the project has no potential impact on their access to other services at the agency. Women who are willing to meet with study staff will be given a referral slip by their providers which they will present to the study research assistant (RA). The study RA will meet potential participants in private locations at each agency. The RA will explain all aspects of the study, including confidentiality and its limits, and address questions. If the participant agrees, they will provide a written informed consent complete the baseline assessment. We will ask participants if they would like us to read consent forms aloud. The RA will emphasize that enrollment in the study is completely voluntary.

Those who consent to participate will be provided with a copy of the informed consent document.

#### The MOSAIC Plus intervention

The MOSAIC Plus intervention will be delivered to those in the intervention arm as specified in the manual, for the duration of 9 months. After completing the screening interviews, the study research assistant will introduce consenting, eligible participants to a MOSAIC Plus trained mentor mother (See training procedures below). The mentor mothers will work as a mentors and advocates in the areas of IPV safety planning, building social support, managing depressive symptoms and enhancing effectiveness obtaining community resources. During the initial phase, mentor mothers will explain their roles as a MOSAIC Plus intervention provider and exchange contact information (i.e., the participant’s contact information and the mentor mother’s study cellphone number). In the following four weeks, the MOSAIC Plus mentors will meet their clients in a private, confidential location to assess if there is ongoing IPV, conduct safety planning, strength and needs assessment. They will collaboratively set a realistic treatment plan for the intervention period. MOSAIC Plus mentor mothers will maintain regular contact with their clients at least once a week (may be more as needed). Information obtained during initial assessment (safety, strengths, needs) will be used to plan the rest of the mentoring intervention in the subsequent months.

After the initial phase is complete, as in the original MOSAIC intervention, mentor mothers will continue meeting their clients (in person or by phone) for a session no less than 60 minutes. This level of contact is high but was feasible and acceptable in the original MOSAIC trial(24). In person meetings will take place in the community at convenient, private, and safe locations (libraries, treatment facilities, public buildings) as suggested by the clients. Telephone meetings will only be made from a study telephone line given to each mentor mother. The mentor mothers will review their clients’ activities since the initial planning meeting and problem-solve any challenges they may encounter. Mentor mothers also facilitate their clients’ access to other services by informing, educating and advocating for them when needed. Their services will involve 1) building social support networks by helping their clients solve any interpersonal difficulties they may have within their networks. Mentor mothers will also work with their clients by linking them with new social support groups and networks available in the community. They will focus on social support because strong social support protects against the negative effects of IPV on mental health (42). 2) Mentor mothers will inform women about services and advocate for their access. Women with experience of IPV may have limited access to information, and hence poor access to services. Mentor mothers will help improve their access to services by actively identifying the needed services, creating proper linkage with providers and advocating for their access by addressing the barriers they may encounter. 3) Plan and monitor their safety by helping their clients create a personalized safety plan and monitor the potential of future violence and address any challenges they may have. Mentor mothers will employ the skills of active listening, reflecting and validating feelings, showing empathy, inspiring hope and facilitating a trusting relationship with other people in their personal networks or people who are service providers. The IPT component in this intervention includes the focus on addressing interpersonal conflict within the woman’s social network. MOSAIC Plus is consistent with the original intent of MOSAIC (support the mother directly), but utilizes a broader conceptualization (i.e., help the mother build her own social support network) and evidence-based IPT strategies for doing so.

#### The control condition

Those in the control condition will receive the MOSAIC intervention without the added IPT component. Participants in the control condition will also receive ongoing monitoring of depressive and PTSD symptoms and suicide ideation with appropriate referrals from mentor mothers. They will also be provided a county-specific IPV resource guide after consenting to enroll in the study.

#### Randomization

Randomization to MOSAIC Plus or MOSAIC interventions will occur after the baseline assessment, in a 1:1 ratio. All baseline assessments are blind. Immediately after randomization, the research assistant will review the study outcome assessment schedule, means of contacting the research staff, and participants’ contacts with all participants. The study statistician will prepare the randomization schedule using opaque sealed envelopes before the enrollment of the first participant.

#### Rationale for choice of target mechanisms

*Social support reduces IPV, depressive, and PTSD symptoms*. IPT’s focus on building social support is a good fit for mothers who have experienced IPV because social support is protective against the negative effects of partner violence (42, 43)and can help reduce future IPV44,45. Perinatal women mothers with ongoing experience of IPV often struggle establishing social networks(43) and therefore additional help developing social support is useful. Social support also helps with depressive and PTSD symptoms(42). Lack of social support is associated with antepartum depressive symptoms(44). Studies suggest that social support, received in the form of informational, instrumental and emotional support, reduces depressive and PTSD symptoms in women with IPV experience(5, 42, 45). Studies in population-based samples of women have found that women with experience of IPV with stronger social support are protected against the negative effects of IPV on mental health including depression and PTSD(42, 46). In contrast, lack of perceived social support is a primary risk factor for developing PTSD following a traumatic experience(47-52). Therefore, social support is a hypothesized target mechanism of the effects of MOSAIC Plus on IPV and mental health outcomes.

**Figure 1:**
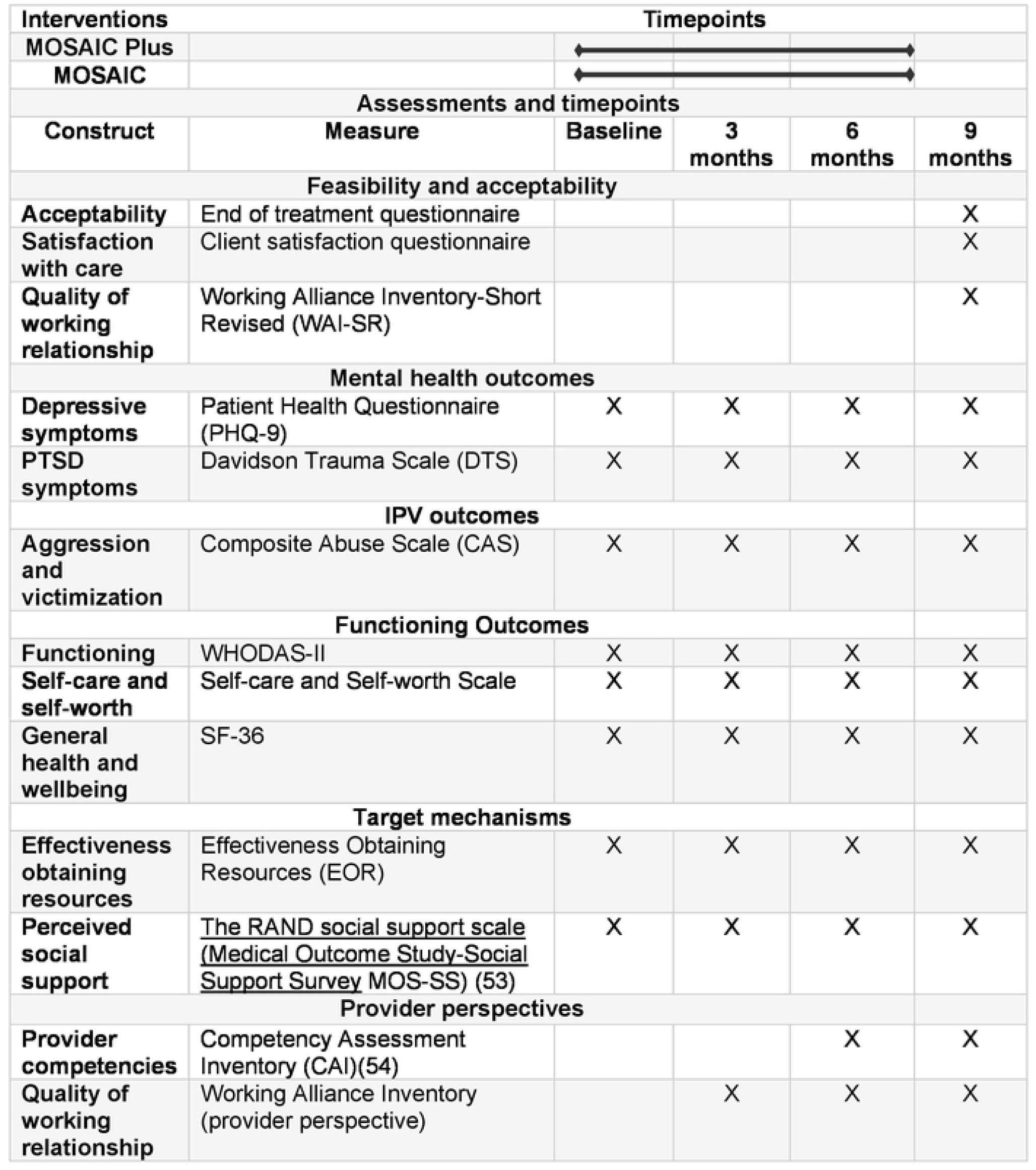
The SPIRIT Schedule

#### Mentor mother training, supervision and supervision

We will recruit 8 mentor mothers and train them to deliver the MOSAIC and the MOSAIC Plus intervention with fidelity. Turnover in community interventionists can be high. Therefore, by training 8 mentor mothers, the study is likely to have at least 6 for the trial. If needed, additional mentor mothers can be recruited and trained. Mentor mothers will be selected with the help of our Community-Based Organization Partners (CBOP), our community partner for this study. With the support from CBOP and its network of partners, we will identify and interview women from Flint Genesee County. Mentor mothers will be: 1) 21 years of age and above, 2) have good listening skills and embody a non-judgmental attitude, 3) a permanent resident in Genesee County, 4) report that they are able to use a computer to upload intervention recordings and have access to a computer with internet, 5) are mothers, 6) have access to reliable transportation and 7) are available up to 2 hours per week by phone or in person. Mentor mothers will have weekly group supervision and case discussion with Dr. Hailemariam. Dr. Hailemariam will also listen to audio recordings of the mentor mother intervention sessions and will provide feedback. During weekly supervision or in the audio files, if a mentor appears to be triggered by a case discussion or talks about being overwhelmed in a group supervision meeting, Dr. Hailemariam will promptly schedule an individual meeting and debrief session with the mentor mother. Mentor mothers will be constantly reminded that if they are feeling overwhelmed by a case (eg., thinking about the case more than they want to, poor sleep, changes in appetite, dreaming about the case, feeling anxious) they will be encouraged to contact Dr. Hailemariam for a debrief and supportive supervision.

##### Safety

We will give strong emphasis to the safety of the mentor mothers. Briefly, we will develop a comprehensive protocol, based on the protocols used successfully in previous MOSAIC studies(24, 53), that covers safety planning with the client and procedures to ensure safety of the mentor mothers. Contents of the protocol will involve leaving the scene if there is an apparent violence and procedures for immediate reporting of any safety concerns and questions. Mentor mothers will be given a study cellphone to communicate with their clients.

They will meet their clients in public places with private space such as health facilities, libraries and parks to reduce potential risk of victimization. To protect mentor mothers’ privacy, mentor mothers will not share their residence addresses and last names with their clients. We will train mentor mothers on how to respond to clients’ substance abuse. Moreover, safety plans will be adapted and tailored to each participating woman based on her needs. Mentor mothers will also be trained on burnout management, self-care and will be offered an opportunity to debrief with study investigators quarterly, and to call on an as needed basis.

##### Fidelity ratings

Intervention adherence and competence ratings will be developed based on the manual and previous IPT adherence and competence scales(30, 54). Adherence rating scales for both conditions will consist of a checklist of tasks to be completed at each meeting. Competence rating scales will be completed from audio recordings of mentor mother visits and will reflect mentor mothers’ general skills, such as reflective listening, safety planning, linking women to services, advocacy and effective goal setting. The study team will use these scales to rate a randomly selected 33% of the open trial tapes. Scale development will continue until the item content is satisfactory and interrater reliability is acceptable (> .80).

**Figure 2:**
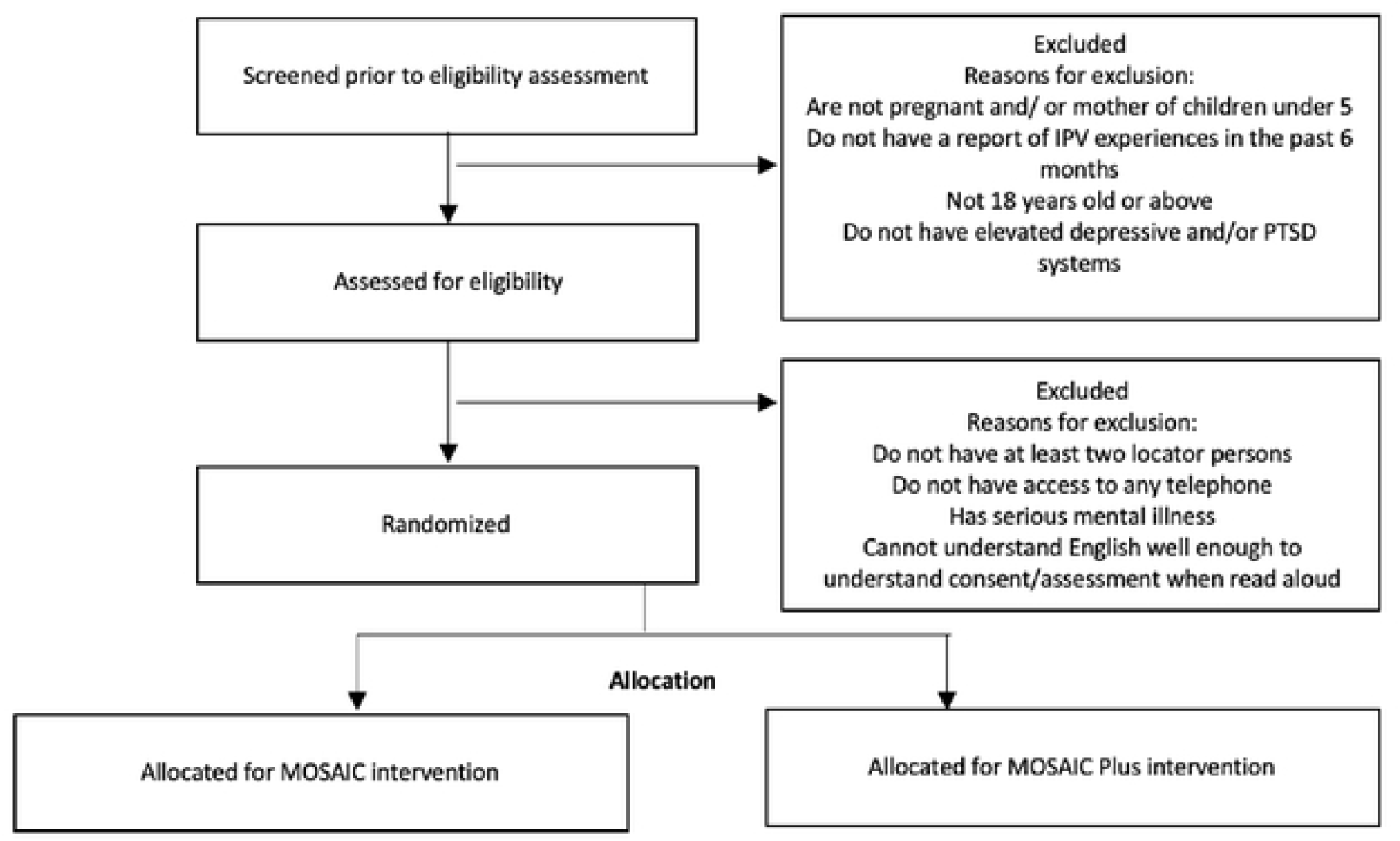
MOSAIC Plus flow chart

##### Assessments

The main goal of this pilot study is to explore the feasibility and acceptability of the MOSAIC Plus intervention. We will also evaluate the feasibility of the research and training procedures. In addition to these main outcomes, we will conduct several assessments to evaluate changes in depressive and PTSD symptoms, reduction in future IPV and changes in effectiveness in obtaining community to resources. The study research assistant (RA) will be trained to fidelity to administer clinical and other assessments. The clinical training process will also be supplemented by standard case vignettes showing elevated depressive and PTSD symptoms to acquaint trainees with common manifestations of IPV, symptoms of depression and PTSD. We will record all RA interviews and do regular quality and fidelity monitoring. Study mentor mothers will not play a role in the data collection process. Data will be collected at baseline, 3 months, 6 months and 9 months (post-intervention completion) (see Table 1: the SPIRIT schedule). We selected the following feasibility, acceptability, and safety metrics based on face validity, clinical experience, and relevant literature.

**Table 1:** Target Outcomes

|  |
| --- |
| **Table 1: Target Outcomes** |

| Assessment method | Target |
| --- | --- |
| **MOSAIC Plus Feasibility and Acceptability ( MOSAIC Plus participants only)** |  |
| Acceptability (End of Treatment Questionnaire) | Endorsing an average score of 3 or more (out of 5) for each rated intervention component). |
| Quality of working relationship (WAI-SR) | Average > 36 (or a mean score of 3 or more [out of 5] on each of the 12 items). We will also descriptively examine goal, tasks, and bond subscales to see where mentor mothers are doing well and less well overall. |
| Satisfaction (Client Satisfaction Questionnaire-8) | Average > 24 (or a mean score of 3 on each of the 8 items) |
| Intervention attendance | 70% of all participants complete at the initial session and at least 2 of 3 booster sessions |
| Intervention Fidelity | MOSAIC Plus intervention achieves at least 80% adherence on a random subset of sessions |
| **Safety (all participants)** |  |
| Adverse events | No serious adverse events or injuries that are possibly, probably, or definitely related to study participation. |
| **Feasibility and Acceptability—Research Procedures (all participants)** |  |
| Recruitment rate | Average of 4-8 enrolled per month |
| MOSAIC Plus intervention procedures | 80% of participants complete MOSAIC Plus intervention sessions at 3 months and 70% at 6-month follow-up |
| Timeliness of assessments | 80% of follow-up assessments occur within 3-weeks of due date |
| Completeness of self-report instruments and interviews | Self-report instruments have 80% of items completed and interviews 90% of items completed in 90% of cases |
| Retention rate | 80% complete 3-mo and 70% complete 6-mo follow up assessments |
| Participant burden | Qualitative responses from exit interviews do not suggest undue burden of intervention or research procedures |

###### Demographic/screening measures

will include age, educational level, marital status, occupation, employment (status, # hours per week), income, race, gestational age/age of children. At follow ups, occupation, employment (status, # hours per week), and income will be repeated. We will also assess relationship information, such as current relationship to the abuser/s and length of relationship. We will use the Composite Abuse Scale (CAS)(37) to include women who report any physical and sexual violence in the past six months. We will also use the Patient Health Questionnaire (PHQ-9) with a cut-off point of ≥ 9(55) and/or the Davidson Trauma Scale (DTS) with a clinical cut off point of ≥ 40 to determine eligibility in terms of depressive and/or PTSD symptoms(39).

###### Primary: Study Feasibility and Treatment Feasibility/Acceptability and safety

We will assess feasibility of the research procedures by examining study recruitment and refusal rates, participants’ willingness to be randomized, follow-up rates, reliability and range of responses to study questionnaires. We will assess the feasibility and acceptability of MOSAIC Plus by examining treatment completion (based on the jointly established treatment plan) and drop-out. We will examine reasons for termination from study and/or intervention for consistent patterns. We will examine the acceptability of the MOSAIC Plus intervention by using data from CTQ-8 treatment satisfaction questionnaire, the End of Treatment Questionnaire, and detailed exit interviews. Clients’ experiences with the mentor mothers, the quality of their working relationship and their level of satisfaction with the service will be evaluated using the Working Alliance Inventory-Short Revised (WAI-SR). Moreover, as in the original MOSAIC study(34), we will also explore perspectives of the mentor mothers regarding acceptability of the MOSAIC Plus intervention. We will use Competency Assessment Inventory (CAI), which tests the attitudes, knowledge, and skills needed to provide high-quality mental health care (i.e., learns and respects clients’ preferences about intervention, creates opportunities for clients to practice skills).

At the end of the open trial and at 2 time-points during the RCT (i.e., after enrolling 20 and 40 participants), we will compute the descriptive data on all target outcomes described in table 2. The scientific team will convene to discuss how our actual outcomes compare to target outcomes. If the team identifies discrepancies between target outcomes and actual outcomes, we will 1) investigate the reason for failure to meet the stated outcomes, and 2) discuss among the research team. Depending on the nature of the discrepancy, we may modify recruitment or assessment procedures, instructions to participants, training procedures, or other trial aspects. Table 2 below presents assessment methods.

**Table 2:** Target Mechanisms

| Table 2: Target Mechanisms |  |
| --- | --- |
| Target Mechanism 1: Social support |  |
| Definition | “Emotional, informational and practical assistance provided in a supportive social network” (69) |
| Evidence that adding IPT approaches increase social support | IPT has been found in many studies to increase social support relative to control conditions(70-73) |
| Evidence that increased social support reduces IPV and depressive and PTSD symptoms | • High social support is associated with lower depressive symptoms(53, 71, 74)<br>• High social support is associated with lower PTSD symptoms, in general and after IPV(50, 75, 76)<br>• Women with IPV who report high social support experience fewer depressive symptoms(46)<br>• Social support protects women from the negative effects of IPV(45, 77) |
| Target Mechanism 2: Effectiveness Obtaining Resources |  |
| Definition | • Women’s ability to access community resources such as legal, healthcare, housing, income assistance, education, jobs, etc.(63) |
| Evidence that IPT increases effectiveness obtaining resources | • IPT increases personal agency to access resources(78). |
| Evidence that effectiveness obtaining community resources reduces depressive symptoms, PTSD symptoms, and subsequent IPV | Access to community resources reduces subsequent IPV(79)<br>Access to community resources reduces depressive symptoms(80)<br>• Access to community resources reduces PTSD symptoms(81, 82) |

##### Clinical and services outcomes

###### Mental health outcomes

Reduction in depressive symptoms will be evaluated using the nine-item Patient Health Questionnaire (PHQ-9), a reliable self-report measure assessing the presence and severity of depressive symptoms(38). We will measure changes in PTSD symptoms by using the 17 item Davidson Trauma Scale (DTS)(39), a reliable and well-validated self-report measure(56) of trauma assessing DSM-5 symptoms of PTSD rated on a five point frequency. We will use the DTS as a cross-sectional measure of PTSD symptoms in three domains including intrusion, avoidance/numbing and hyperarousal. Respondents will be asked to identify the most troubling traumatic experience in the past week and measure its severity and frequency in 5-point scales.

##### IPV outcomes

We will use the Composite Abuse Scale to measure victimization in the context of romantic and partner relationship for each intimate relationship over a specified period of time. The CAS has 4 dimensions including severe combined abuse, emotional abuse, physical abuse, and harassment.

##### Functioning outcomes

The 12-item WHO-Disability Assessment Schedule (WHO-DAS-II)(57) will be used to measure *functioning* in the domains of cognition, mobility, self-care, getting along/interaction with other people, life activities and participation in community activities. A nine-item assessment tool measuring Self-care and Self-worth will be used assess changes in self-care. The scale was developed based on our team’s previous qualitative work with high-risk women(58). Items included “I take good care of myself,” “I need to take care of people around me before I take care of myself,” “I am worth taking care of,” “I am worth protecting,” “I deserve to be able to take care of myself,” “I deserve to protect myself,” “I neglect myself,” “I put myself in dangerous situations,” and “I take care of my needs,” and were answered on a 5-point Likert Scale from “Strongly agree” to “Strongly disagree.” The scale has an internal consistency reliability (Cronbach’s alpha) of .77. To measure *general health and wellbeing*, we will use SF-36 (59). SF-36 has a good internal validity in research, community and clinical settings (59, 60).

###### Target mechanisms

We will measure *social support* using the RAND social support survey instrument (also known as Medical Outcome Study-Social Support Survey (MOS-SS)(61) that measures functional social support covering multiple dimensions including emotional/informational, tangible, affectionate and social interaction. The scale has previously been used in a study of maternal depression(62). We will use the Effectiveness Obtaining Resources Scale (EOR)(63) to assess IPV exposed perinatal women’s’ effectiveness in obtaining resources from formal sources in 13 areas: housing, material, goods and services, education, employment, health care for themselves and their children, child care, transportation, social support, legal assistance, financial issues, and other issues regarding themselves and their children. The EOR was developed for use among women with IPV.(63) It is reliable, valid, and shows significant changes over time in clinical samples (63-65).

##### Provider perspectives

We will use Competency Assessment Inventory (CAI) to evaluate the skills, attitude and knowledge needed to deliver a high quality mentoring intervention(66). We will also use the provider version of Working Alliance Inventory(67) to assess mentor mothers’ perceptions of agreement on mentorship goals, agreement on mentorship tasks, and the quality of the mentoring relationship with each participant mother.

## Data Analysis

As this is an intervention development study, our primary goal is assessment of feasibility and acceptability of the MOSAIC Plus intervention and research procedures. However, pilot data can be used to demonstrate whether the effects of treatment look promising across a set of outcome variables, to begin to examine distribution of outcome variables to inform future analytic strategies, and to suggest, in concert with results from larger scale clinical trials in related fields, the range of effect sizes that would be reasonable to expect in a future trial. We will obtain the between treatment condition effect size estimates (with 95% CI) at each assessment (e.g., Cohen’s d or h) and the correlation between the same dependent variable at adjacent assessments. We will have complete data on approximately 40 participants (the minimum expected after participant loss due to attrition). Sample size guidelines for treatment development from Rounsaville et al. (68) recommend 15 to 30 participants per cell. Given that group means begin to stabilize by around 15, we believe a sample of 20 in each condition should provide some information relevant to demonstrating potential promise for MOSAIC Plus.

### Mental Health Outcomes

We will: (1) Calculate the effect size and 95% CI for reduction in depressive symptoms (PHQ-9 scores). Exploratory tests for differences between conditions will use HLM, with baseline scores as a covariate. (2) Calculate the effect size and 95% CI separately for reduction in PTSD symptoms (DTS scores) using HLM with baseline DTS score as a covariate.

### IPV outcomes

We will calculate the effect size and 95% CI separately for reduction in CAS scores using HLM with baseline CAS score as a covariate.

### Functioning outcomes

For secondary outcomes, separate calculations of the effect sizes and 95% CI will be performed. These include improvement in functioning (WHODAS-12) self-care (Self-care and Self-worth scale) and general health and wellbeing (SF-36). Separate exploratory tests for differences between conditions will use HLM, with baseline scores as covariates.

### Target mechanisms

We will separately calculate the effect size and 95% CI for the effect of the intervention on proposed target mechanisms including improved social support (measured using MOS-SS) and effectiveness obtaining resources (measured using EOR scale). Separate exploratory tests for differences between conditions will use HLM, with baseline scores as covariates. We will then explore the association of each of these target mechanisms with changes in our primary outcome (reduction in depressive and PTSD symptoms) from baseline through 9 months. These exploratory analyses will inform full tests of mediation (i.e., tests of the hypothesis that the effects of MOSAIC Plus on reduced depressive symptoms is mediated through improvement in social support and effectiveness obtaining resources) in a subsequent fully powered trial. Although we expect the MOSAIC Plus intervention show evidence of preliminary effectiveness, should the intervention show no or limited evidence, exploratory tests of target mechanisms will also provide some initial information about whether MOSAIC Plus’ limited effectiveness was due to failure to engage target mechanisms or to the target mechanisms not being associated with the final outcome. Target mechanisms are presented in table 2 below.

### Personalization and processes

We will explore marital status, maternal age, race/ethnicity, household income, parity, severity of IPV experience, gestational age at enrollment, and number of lifetime trauma experiences (as assessed by the Trauma History Questionnaire) (83) as predictors and moderators of treatment outcome. These variables were also used in the original MOSAIC study(24) and other studies with women with IPV experiences(25, 46, 84). We expect that MOSAIC Plus will be appropriate for a full range of perinatal women with IPV experience. Preliminary analyses of dose-response effects will also be conducted.

### Treatment Integrity

We will calculate scale reliabilities of adherence and competence ratings using both individual item correlations and total intraclass correlations. We will compute scale validity by correlating adherence and competence ratings to intervention outcomes, to each other, and to expert (MH and JEJ) ratings. Adherence and competence ratings of trained raters will be compared to expert global ratings to determine cut-off scores with sufficient sensitivity and specificity.

## Ethics and dissemination

The MOSAIC (MOthers’ AdvocateS In the Community) for Pregnant Women and Mothers of Children Under 5 With Experience of Intimate Partner Violence (MOSAIC Plus) study was approved by Michigan State University Biomedical IRB (#00 512720). All study staff also completed the Michigan State University Human Subject training certificate and good clinical practice (GCP) trainings. The study RA will meet potential participants in private locations at each agency. The study research assistant will explain all aspects of the study to the participants and a written informed consent will be obtained. We will ask participants if they would like us to read consent forms aloud. The research assistant will emphasize that enrollment in the study is completely voluntary. Those who consent to participate will be provided with a copy of the informed consent document. The study was also registered in www.clinicaltrials.gov under identifier # NCT05106361, date of registration 03 November 2021, https://clinicaltrials.gov/ct2/show/NCT05106361.

### Dissemination and data sharing

The research outcomes generated from this R34 will be disseminated in a timely fashion. The study investigators are committed to reporting the study results both in national and international platforms per the proposed timeline of the study. After data have been collected and study results published, de-identified electronic data will be made available to other qualified researchers upon request. The request will be evaluated by the investigators to ensure that it meets reasonable standards of scientific integrity. For rapid, responsible and broad data sharing, we will follow the standard NIH-funded clinical trial data sharing procedures. We will also ensure compliance of our data sharing and reporting procedures with the Michigan State University’s internal policy regarding clinical trials registration and reporting results. We will place the de-identified dataset, along with the data dictionary and documentation of data collected, into the NIMH Limited Access Dataset Repository. We will submit primary results for publication by the end of the project period, and will have final de-identified datasets and data dictionaries available by CD and on the NIMH Limited Access Dataset Repository within required timeframes. We are committed to doing research that will change clinical practice. We will also work with our other community partners to share results of the study with their state and national networks, through presentation at their conferences and meetings, newsletters, flyers, and other strategies that they deem to be appropriate.

## Discussion

The current study is designed to provide evidence for the feasibility and acceptability of a mentor mother-delivered intervention for IPV and depressive and/or PTSD symptoms among pregnant women and mothers with children under the age of 5. Moreover, the study also aims to pilot research procedures for a subsequent fully powered trial comparing MOSAIC Plus to the original MOSAIC.

Addressing IPV and its associated mental health symptoms (i.e. depression and PTSD) is important because unaddressed mental health challenges and IPV increase the woman’s risk of mortality, morbidity, future child abuse, suicidal ideation, and femicide(84). Women with elevated depressive and PTSD symptoms are at greater risk of experiencing future IPV(3, 85). Moreover, the stigma of mental illness also reinforces abusers’ ability to manipulate, control and discredit survivors, and weaken vital social support(85). Therefore, depressive and PTSD symptoms not only follow IPV and are associated with suffering and morbidity, they potentiate future IPV risk. This bidirectional relationship between depression/PTSD and IPV suggests that addressing these issues simultaneously may help end the vicious cycle of victimization and mental health vulnerability. Therefore, an integrated intervention for reducing subsequent IPV, elevated symptoms of maternal depression and PTSD in pregnant women and mothers with children under 5 is needed.

Evidence exists that paraprofessionals can deliver mental health interventions for perinatal women with similar fidelity to mental health professionals(86). Results from the original MOSAIC study also confirm that paraprofessional/mentor mother-delivered interventions reduce the risk of subsequent IPV among pregnant women and mothers with children under 5(24).

IPT can be delivered by non-professional lay providers. Community-based, non-professional delivered, evidence-based mental health interventions can help reduce health disparities present in underserved groups(87). For example, IPT has been found to be effective for treating major depressive disorder using providers with no particular educational background, who were native Ugandans treating individuals in rural villages (88). Several other studies have also shown that non-professional or paraprofessional workers with appropriate clinical supervision can effectively deliver IPT (54, 89, 90), including studies conducted by our team(54, 89). Therefore, incorporating IPT principles into MOSAIC for use by the mentor mothers is feasible and appropriate.

To date, no study has found that an IPV focused intervention for any group of women significantly reduced both future IPV risk and depressive and PTSD symptoms. Addressing depressive and PTSD symptoms while effectively reducing future risk of IPV in mothers who report IPV is crucial.

This study expands the literature because it will develop and pilot-test a paraprofessional-delivered intervention to address symptoms of maternal depression and PTSD in a highly vulnerable sample; pregnant women and mothers with young children who report experience of IPV.

## Data Availability

No datasets were generated or analysed during the current study. All relevant data from this study will be made available upon study completion.

## Declarations

### Consent for publication

N/A

### Availability of data and materials

N/A

### Authors’ contributions

MH is the principal investigator of the study. JJ and CZ are co-investigators. AT is the consultant and original creator of the MOSAIC intervention. MH drafted the manuscript. JJ, CZ and AT reviewed and contributed to different parts of the manuscript and approved it.

## Acknowledgements

N/A

